# A Systematic Review of effect of Non-Invasive Brain Stimulation on Cognition Impairment after a Stroke and Traumatic Brain Injury

**DOI:** 10.1101/2020.03.06.20032243

**Authors:** Takatoshi Hara, Aturan Shanmugalingam, Amanda McIntyre, Amer M. Burhan

## Abstract

**Background:** In recent years, the potential of non-invasive brain stimulation (NIBS) for therapeutic effects on cognitive functions has been explored for stroke and traumatic brain injury (TBI) populations.

**Methods:** All English articles from the following sources were searched from inception up to December 31, 2018: PubMed, Scopus, CINAHL, Embase, PsycINFO and CENTRAL. Randomized and prospective controlled trials, including cross-over studies, were included for analysis. Studies with at least five individuals post stroke or TBI, whereby at least five sessions of NIBS were provided and used standardized neuropsychological measurement of cognition, were included.

**Results:** A total of 17 studies met eligibility criteria which included 546 patients receiving either repetitive transcranial magnetic stimulation (rTMS) or transcranial direct current stimulation (tDCS). Sample sizes ranged 5-25 subjects per group. Seven studies used rTMS and ten studies used tDCS. Target symptoms included global cognition (n=8), memory (n=1), attention (n=1), and unilateral spatial neglect (USN) (n=7). Nine studies combined rehabilitation or additional therapy with NIBS. Six of ten studies showed significant improvement in attention, memory, working memory, and executive function. In the USN study, five of the seven studies had a significant improvement in the intervention group.

**Conclusions:** The effect of NIBS on executive functions including attention and memory post stroke or TBI yielded mixed results with variable stimulation parameters. A significant, consistent improvement was observed for USN post stroke or TBI. Future studies using advanced neurophysiological and neuroimaging tools to allow network-based approach to NIBS for cognitive symptoms post stroke or TBI are warranted.

## INTRODUCTION

After a stroke and traumatic brain injury (TBI), cognitive impairment may lead to significant dysfunction for individuals. The main symptoms are memory disorder, attention disorder, executive dysfunction, and social behavior disorder.^1^ In a national epidemiological cohort study of population and prevalence after brain injury in the chronic phase, Nakajima et al.^1^ reported that the most common cognitive symptoms were memory impairment (90%), attention disorder (82%), and executive function impairment (75%). The scope and severity of cognitive symptoms depend on many factors, including injury mechanism, demographic and social factors. According to previous studies, impairment of attention disorder occurs among 42-92% of individuals in the acute phase and is evident in 24-51% at discharge from acute care.^2,3^ Studies have reported that memory impairment persists in 23-55% of individuals up to 3 months post stroke and 11-31% at one year post stroke; this figure is similar in the TBI population (25%).^4,5^ Unilateral Spatial neglect (USN) is an attention disorder that occurs after stroke and is characterized by the inability to orient or respond to or report the stimuli appearing contralateral to lesion side.^6^ The incidence of USN has been reported as low as 8% and as high as 90% in stroke patients.^7^ These cognitive issues cause significant limitation in rehabilitation effort, resuming work, and the need for assistance.^8,9^

The basic focus of cognitive rehabilitation is directed towards achieving changes that improve the person’s everyday functioning. ^10^ Cognitive rehabilitation is thought to have little direct effect on cognitive domains, rather, acquisition of compensatory tools and strategy is fundamental to cognitive rehabilitation.^11^ There are several systematic reviews examining cognitive rehabilitation after a stroke or TBI.^7,12-14^

Recently, the role of non-invasive brain stimulation (NIBS) in rehabilitation post stroke and TBI has attracted significant attention.^15^ In general, NIBS techniques use electrical and/or magnetic energy to induce change in excitability of the underlying brain cortex in a non-invasive fashion and potentially induce long-lasting neuroplastic changes. Many stimulation protocols have been developed, which can provide excitatory or inhibitory stimuli to the cerebral cortex.^16,17^ This has resulted in therapeutic applications of NIBS to treat depression.^18,19^ In recent years, the potential of NIBS to have therapeutic effects on cognitive functions has been explored for stroke and TBI populations as well.^20-22^ We have previously reported a case in which improvement was achieved by using rTMS combined with intensive rehabilitation in combination with higher brain dysfunction following brain injury.

Furthermore, the use of single photon emission computer tomography demonstrated changes in perfusion in the rTMS target sites and areas surrounding the targets.^21^ In our case studies, the stimulation parameters were individually selected based on clinical symptoms and pre-intervention neuroimaging. Rehabilitation of complex cognitive symptoms post stroke and TBI impose significant challenges in identifying stimulation parameters including stimulation site, stimulus characteristics, frequency and duration of stimulation. Based on the increasing number of studies in this novel field, we aimed to perform a systematic review to investigate the effect of NIBS in the rehabilitation of cognitive impairment after stroke and TBI.

## MATERIALS AND METHODS

### Literature Search Strategy

The following sources were searched from inception and up to December 31, 2018 for literature published in the English language: PubMed, Scopus, CINAHL, Embase, PsycINFO and CENTRAL. Selected keywords included Acquired brain injury,Traumatic brain injury, Brain injury, Head injury, Craniocerebral trauma, Stroke, Cerebral Vascular Accident, Ischemic Stroke, Hemorrhagic Stroke, Non-invasive brain stimulation, Transcranial magnetic stimulation, Theta-burst stimulation, Quadripluse stimulation, Transcranial Electrical Stimulation, Transcranial direct-current stimulation, Transcranial Alternating current stimulation, Cognition, Memory, Attention, Unilateral spatial neglect, Executive functioning. Variations of keywords were individualized for each scientific database. All retrieved articles were reviewed to ensure all relevant articles were included for data synthesis. The PubMed search strategy is illustrated in Appendix 1.

### Study Selection

Articles reporting on randomized and prospective controlled trials (RCT and PCT, respectively), including cross-over studies, were included for review. We included studies in which NIBS was used for cognitive rehabilitation post stroke and/or TBI, reported cognitive function pre- and post-intervention and included a minimum of five daily sessions of NIBS. Articles reporting on protocols, retrospective studies or case reports were excluded. We included studies reporting on at least five patients, who were 18-85 years old post stroke or TBI. Two authors (TH and AS) independently reviewed all potential studies for inclusion against the eligibility criteria. They examined the title and abstract and, where necessary, the full text of studies to assess if they were eligible for inclusion. If they could not reach agreement by discussion, a third author (AB) made the final decision about eligibility.

### Date Extraction

Two authors (TH and AS) independently used a standard form to extract study characteristics and outcome data from the studies. Discrepancies were checked against the original data. A third author (AB) made the final decision in the cases of disagreement. Data extracted from each study included author, year, sample size, sex, age, time between onset and treatment, target symptom, stimulation site, each NIBS parameter, outcome measures, and results..

### Methodological Quality

We assessed the methodological quality of selected studies as described in the Physiotherapy Evidence Database (PEDro) scoring system.^23^ This assessment has 11 items on study quality that are answered with yes (score=1) or no (score=0). The first item is a measure of external validity and is not used in calculating the final score. Based on this assessment, all studies were given a level of evidence according to a modified Sackett Scale.^24^

## RESULTS

### Study Selection

After screening 961 citations, 36 potentially relevant studies were identified. After reviewing the full text, 17 studies met the predetermined inclusion criteria (Figure 1). Among the excluded articles, 10 had less than 5 sessions or 5 days of NIBS. In 5 studies the outcomes did not include neuropsychological testing, 2 studies had less than 5 patients, 1 article was only a protocol description, and 1 article was comparative study that classified the stimulation methods according to the paralyzed side and compared the change of attention assessment.

**Figure 1.**
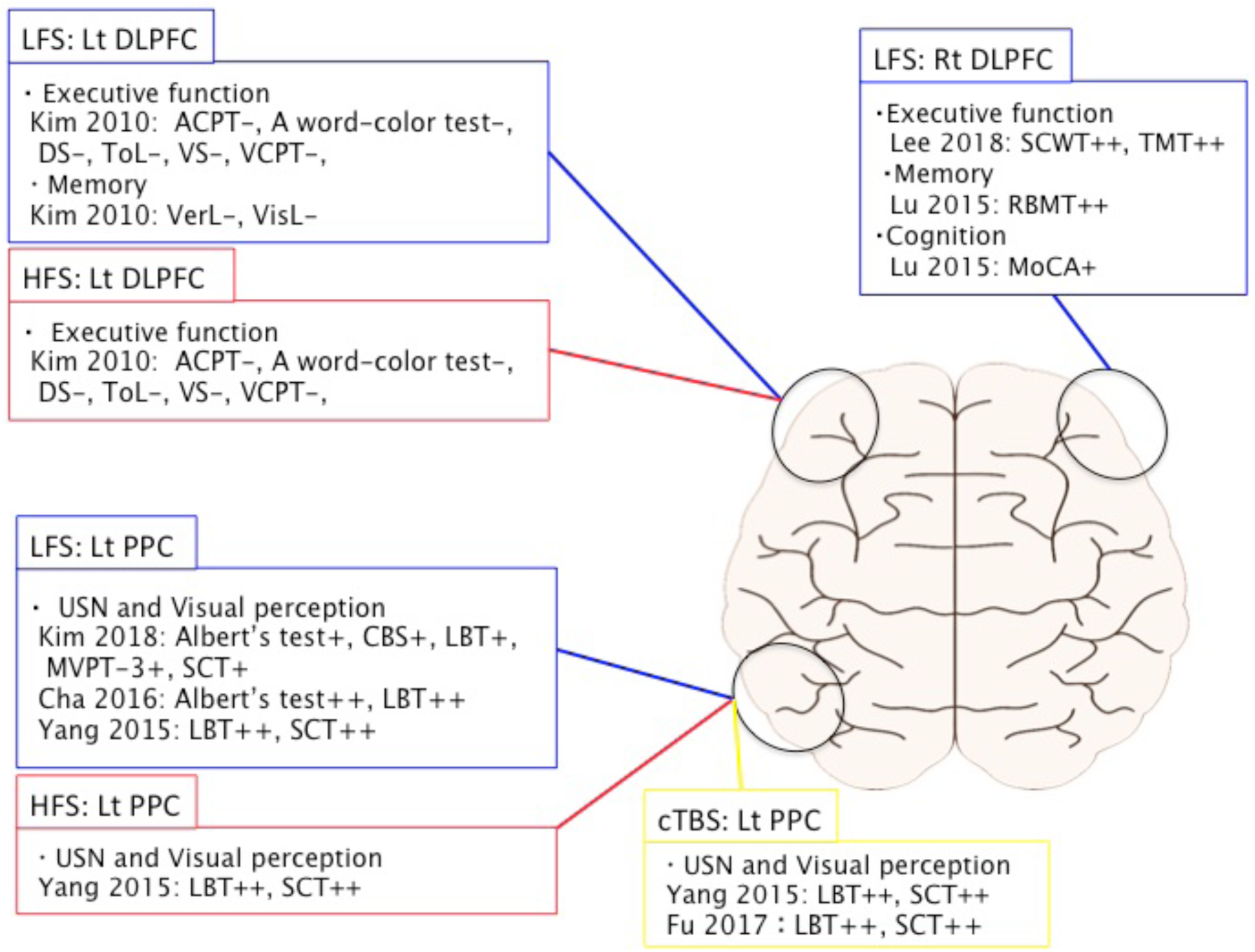
Relationship between stimulation site and neuropsychological test results in rTMS: *Note:* ++ = a significant improvement before and after the intervention and a significant improvement compared to the control group. + = significant improvement before and after the intervention; however, no significant improvement compared to the control group. - = no significant improvement before and after the intervention. *Abbreviations:* DLPFC=Dorsolateral prefrontal cortex, PPC=Posterior parietal cortex, LFS=Low frequency stimulation, HFS=High frequency stimulation, cTBS= continuous theta-burst stimulation, ACPT=Auditory continuous performance test, CBS=Catherine Bergego Scale, DS=Digit Span, LBT=Line bisection test MoCA=Montreal Cognitive Assessment, MVPT= Motor-Free Visual Perception Test, RBMT=Rivermead Behavior Memory Test, SCT=Star cancellation test, SCWT=The Stroop Color Word Test, TMT=Trail Making Test, ToL=Tower of London test, VCPT=visual continuous performance test, VerL=verbal learning test, VisL=visual learning test, VS=Visual span

### Study Characteristics

The details of each study are provided in Table 1. In terms of study design, 15 articles in this review were RCTs, with the remaining two articles PCTs with cross over design. The pooled sample size was 546 individuals who received repetitive transcranial magnetic stimulation (rTMS) or transcranial direct current stimulation (tDCS) with a sample size varying from 5 to 25 subjects per group. While 13 studies included individuals with stroke, just 4 studies included individuals with TBI. The age range of the intervention group was 29.2-70.3 years, and for the control group, 28.2-70.6 years. The time between onset and treatment ranged 19.2 days to 4.8 years (one study was unclear). Twelve studies were ranked as Level 1 evidence and five studies as Level 2 evidence. In the majority of studies (88.2%), subjects were randomly allocated to groups appropriately. With the exception of two studies, intervention and control groups were similar at the baselines regarding the most important prognostic indicators. Blinding was highly variable among studies. A total of 88.2% of studies yielded at least one important outcome measure from more than 85% of the subjects initially assigned to a group. In addition, the results of statistical comparisons between groups and the presentation of point measures and measures of variability were adequately performed in many studies (70.5%, 94%).

**Table 1.**
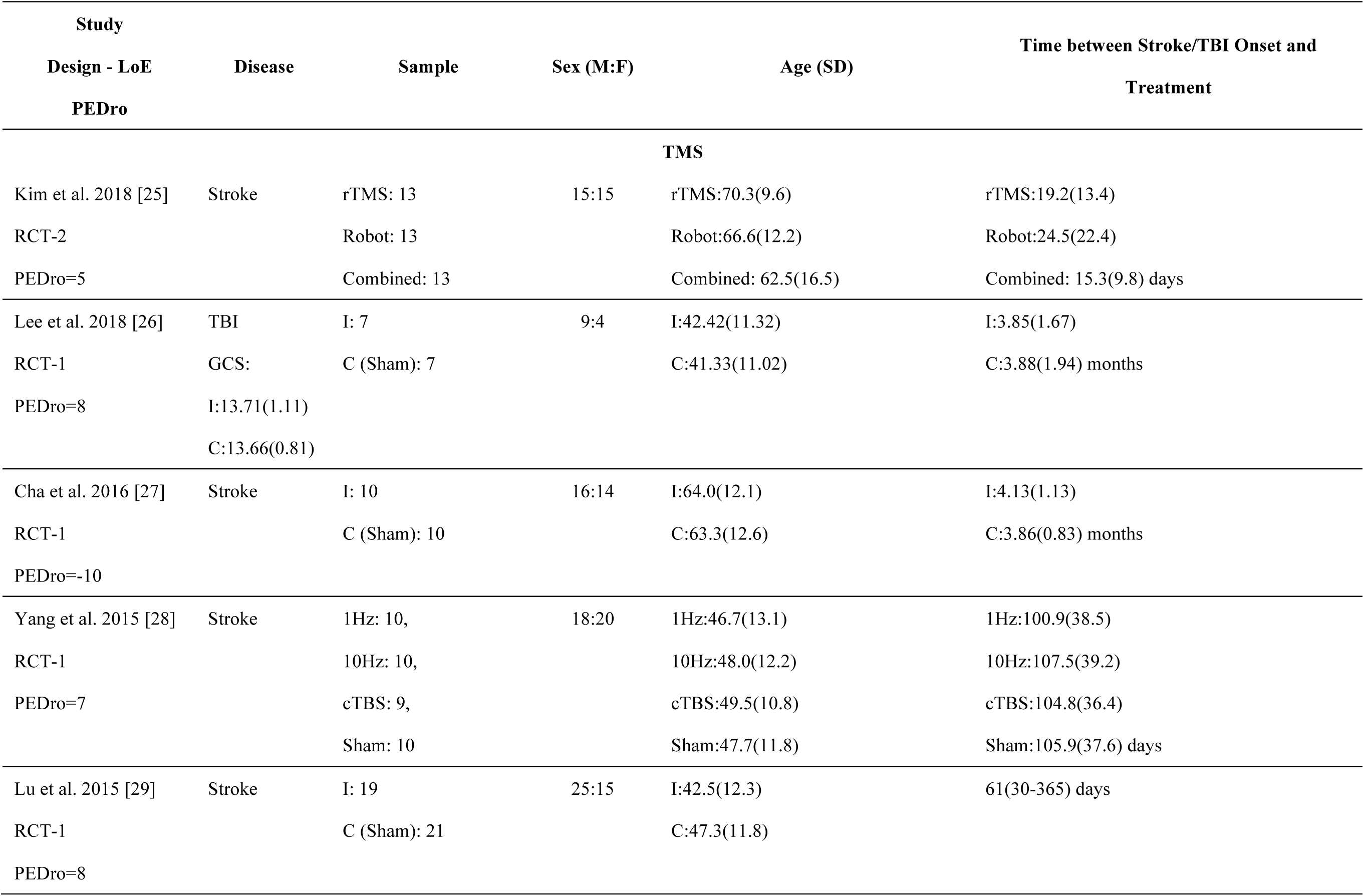
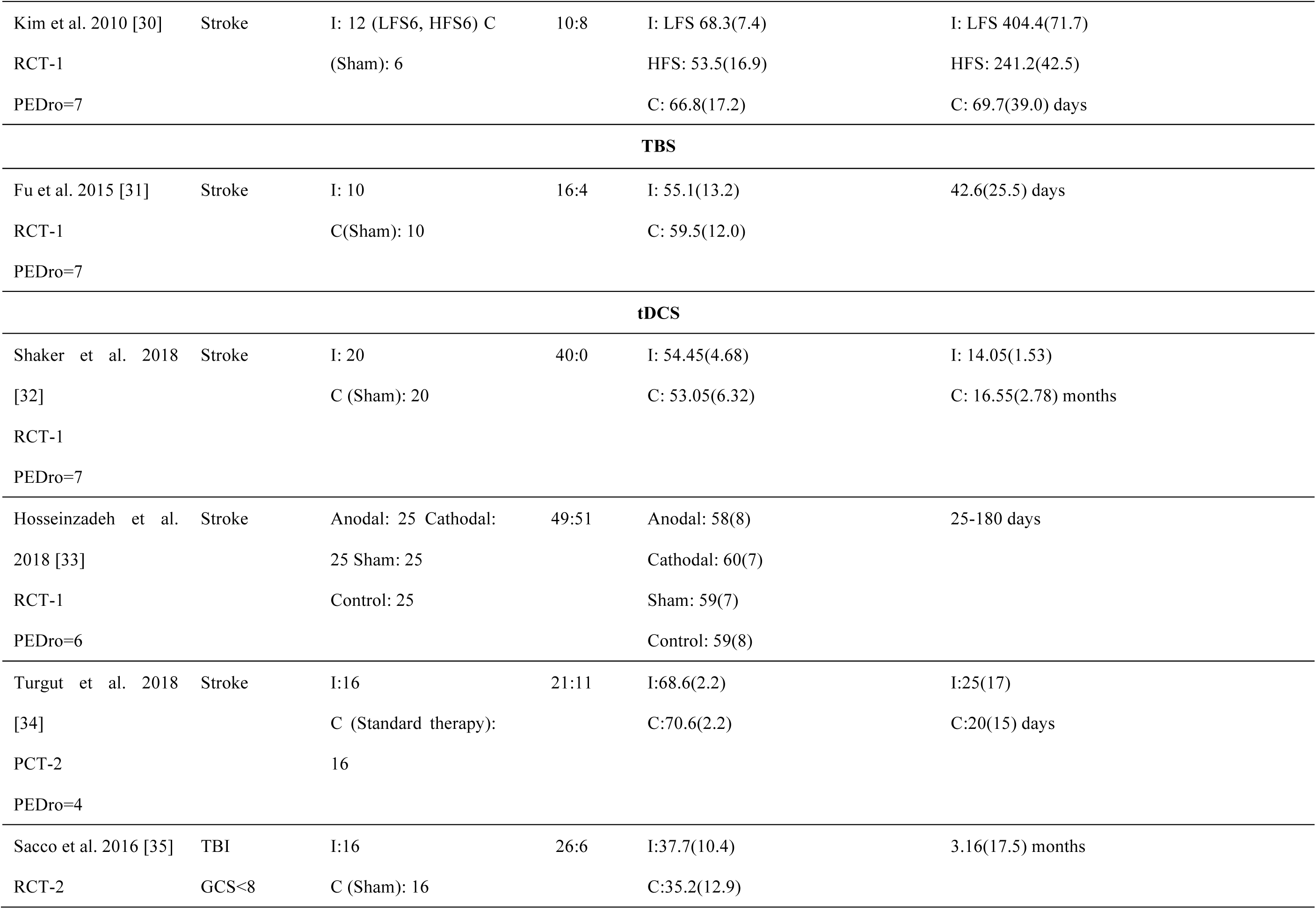
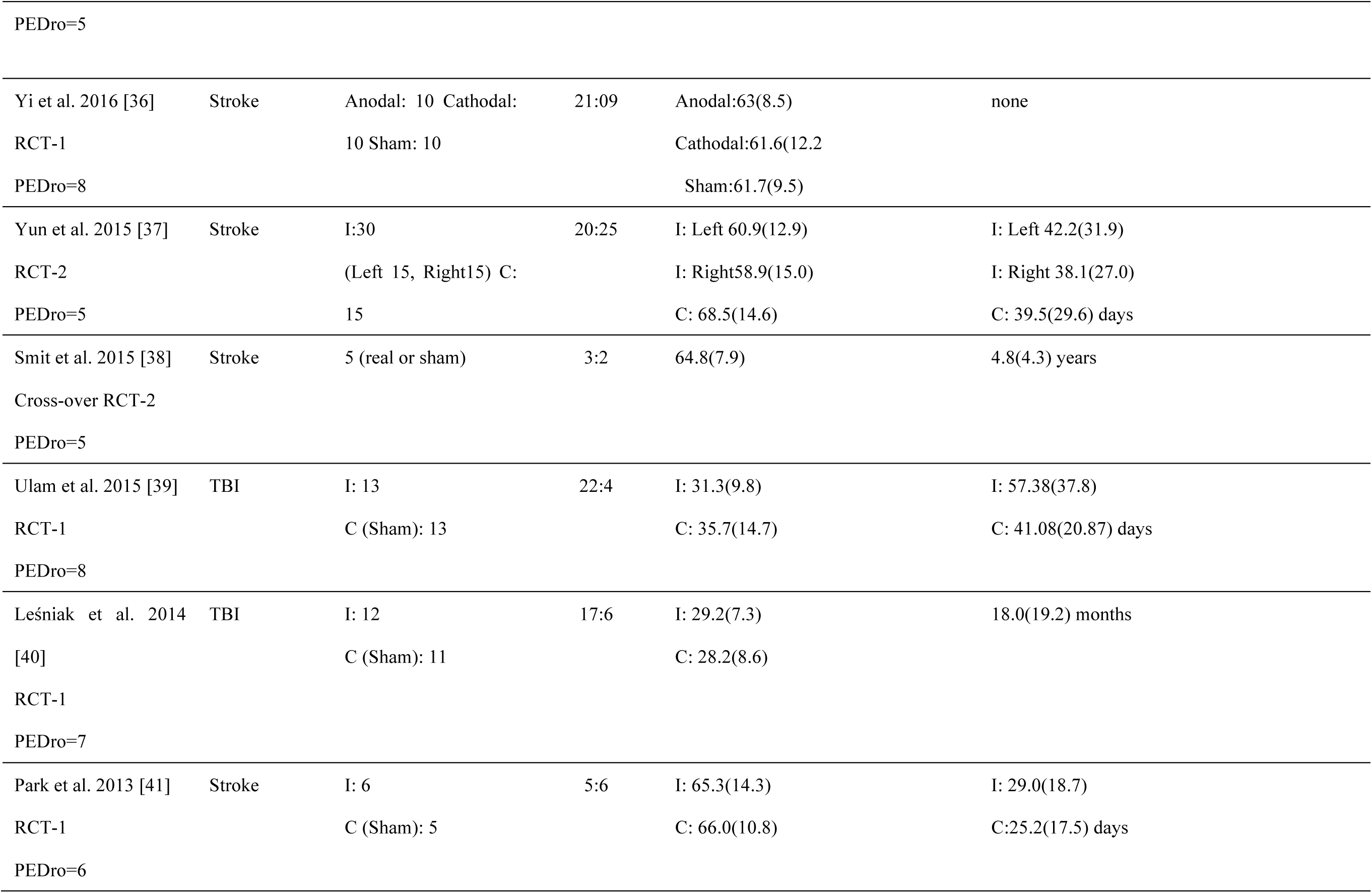
Study Characteristics

### Treatment Characteristics, Outcomes, and Results

The treatment characteristics, outcomes, and results for each study are listed in Table 2. There were seven studies using rTMS and ten studies using tDCS. In total, 14 studies combined rehabilitation or additional therapy with NIBS. In terms of the impairment assessed, eight articles reported on global cognition and seven on USN. Kim et al.^25^ compared rTMS alone, robot rehabilitation, and combination therapy. In terms of the stimulation pattern, five articles of rTMS study used inhibition stimulation pattern. Yang et al. ^28^ used 1Hz, 10Hz, and continuous theta-burst stimulation (cTBS). Kim et al.^30^ used 1Hz and 10Hz of rTMS. Eight articles studied tDCS using anodal simulation. Hosseinzadeh et al. used anodal tDCS for the left Superior temporal gyrus (STG) and cathodal tDCS for the right Posterior Parietal Cortex (PPC). Similarly, Yi et al.^36^ used anodal tDCS for the right PPC and cathodal tDCS for the left PPC. Only eight articles performed a follow-up assessment after stimulation.

**Table 2.**
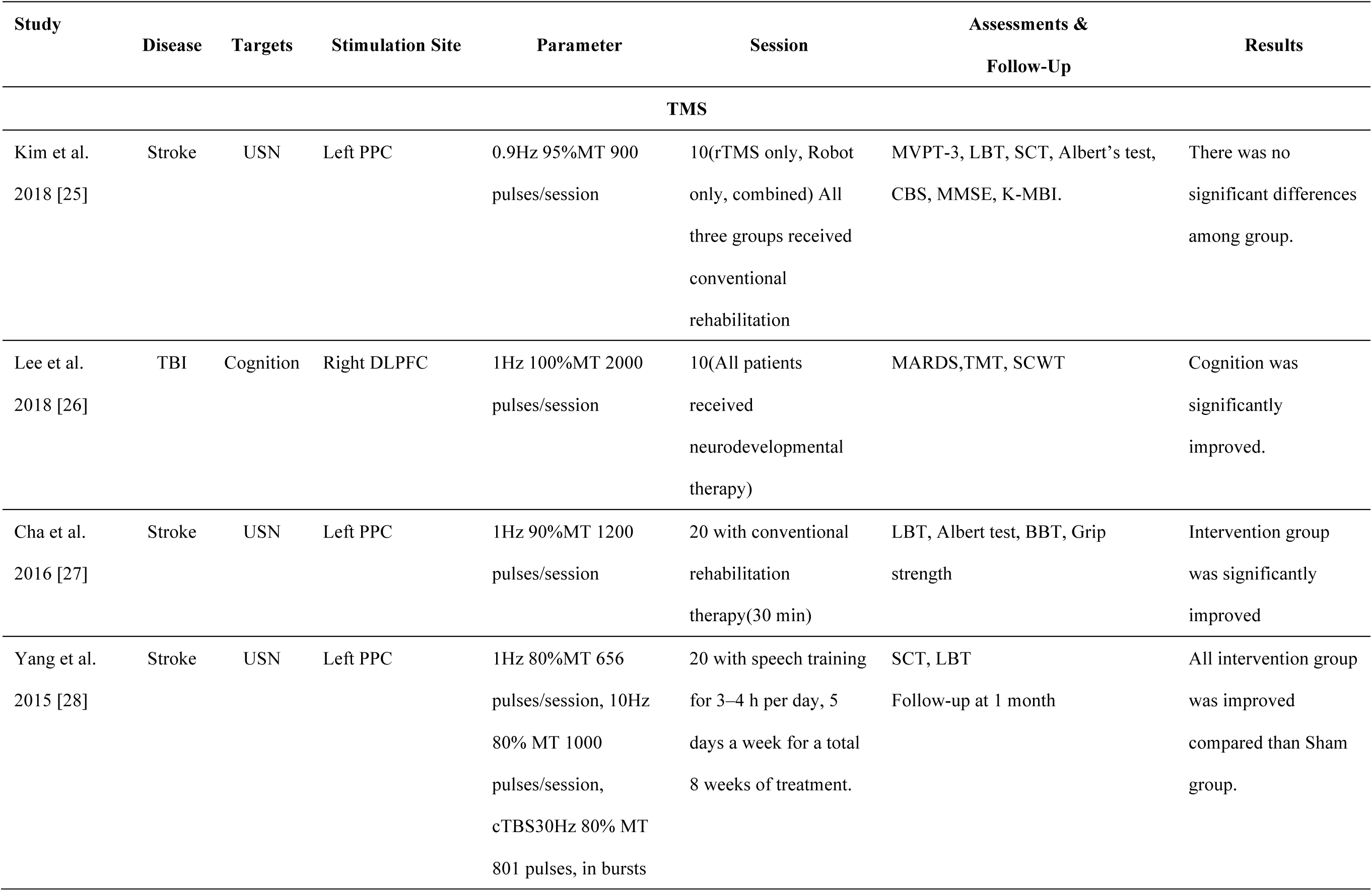
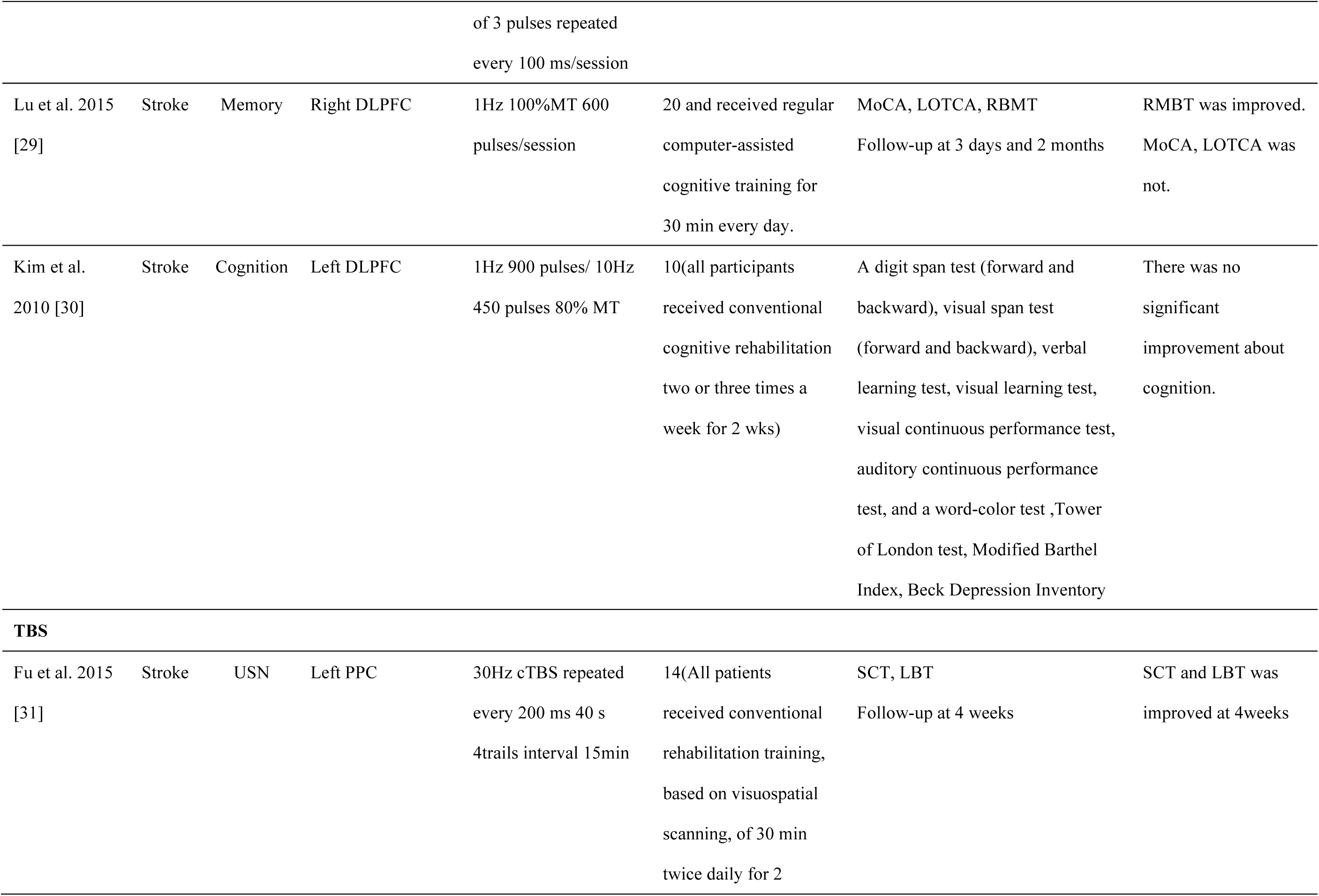
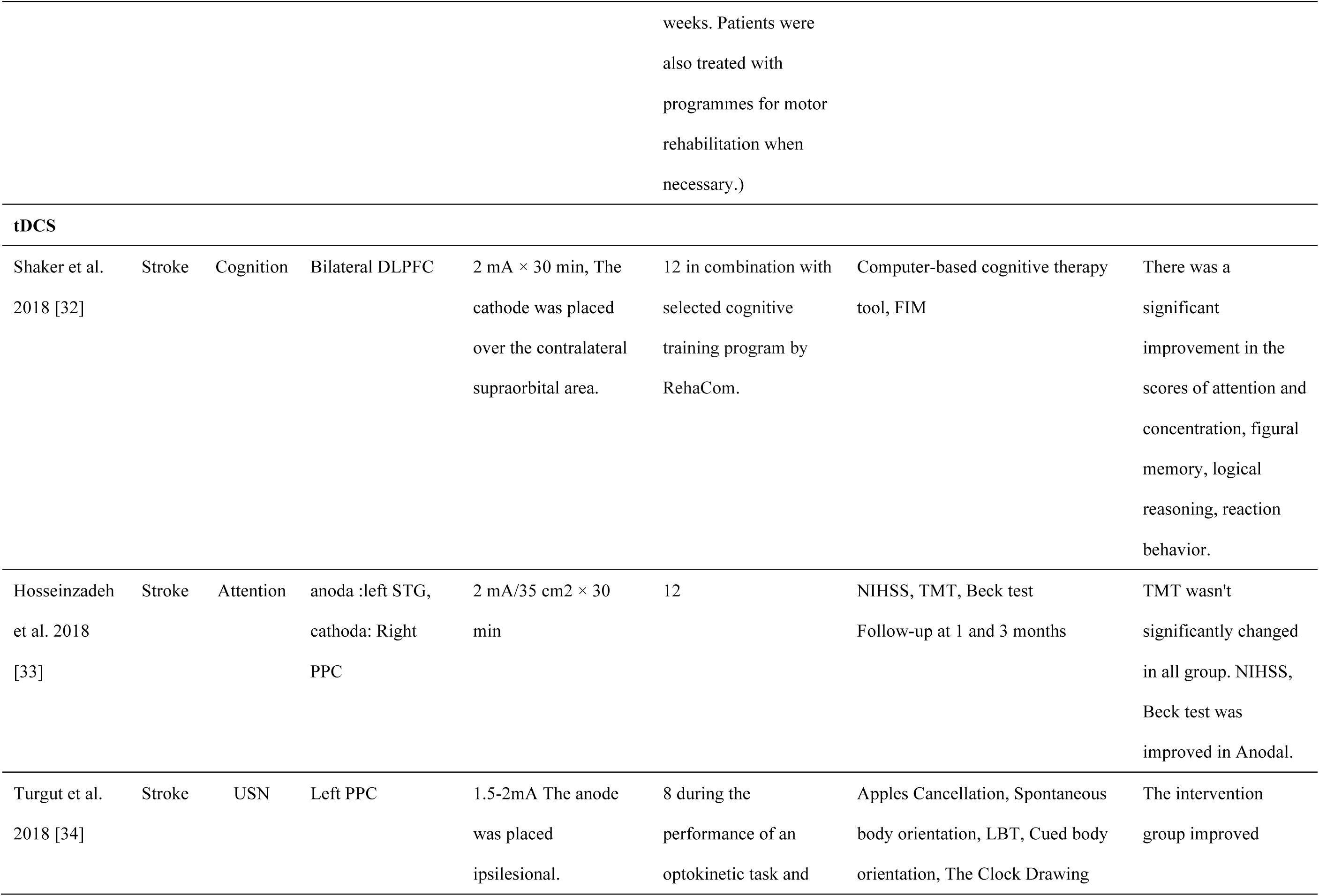
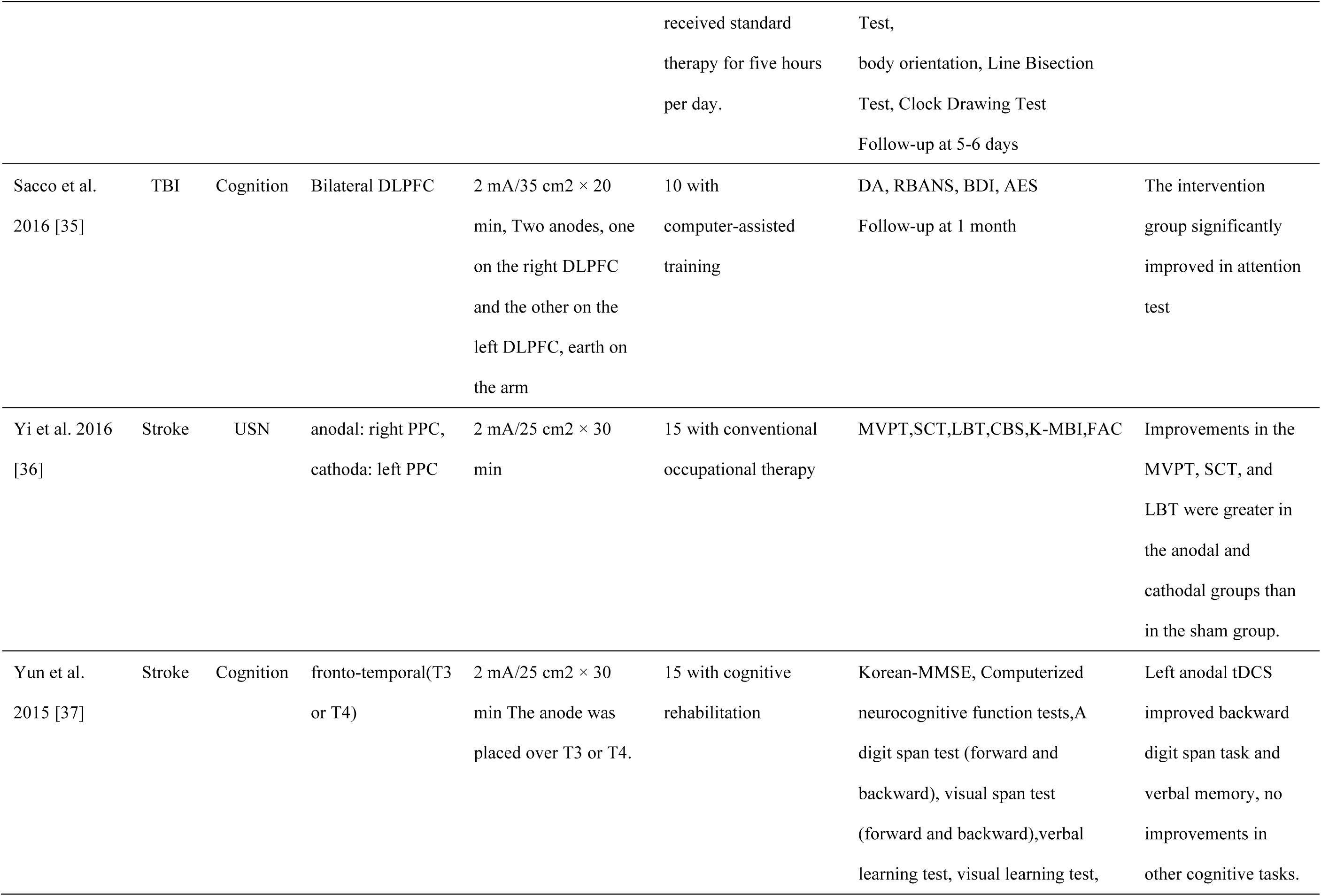
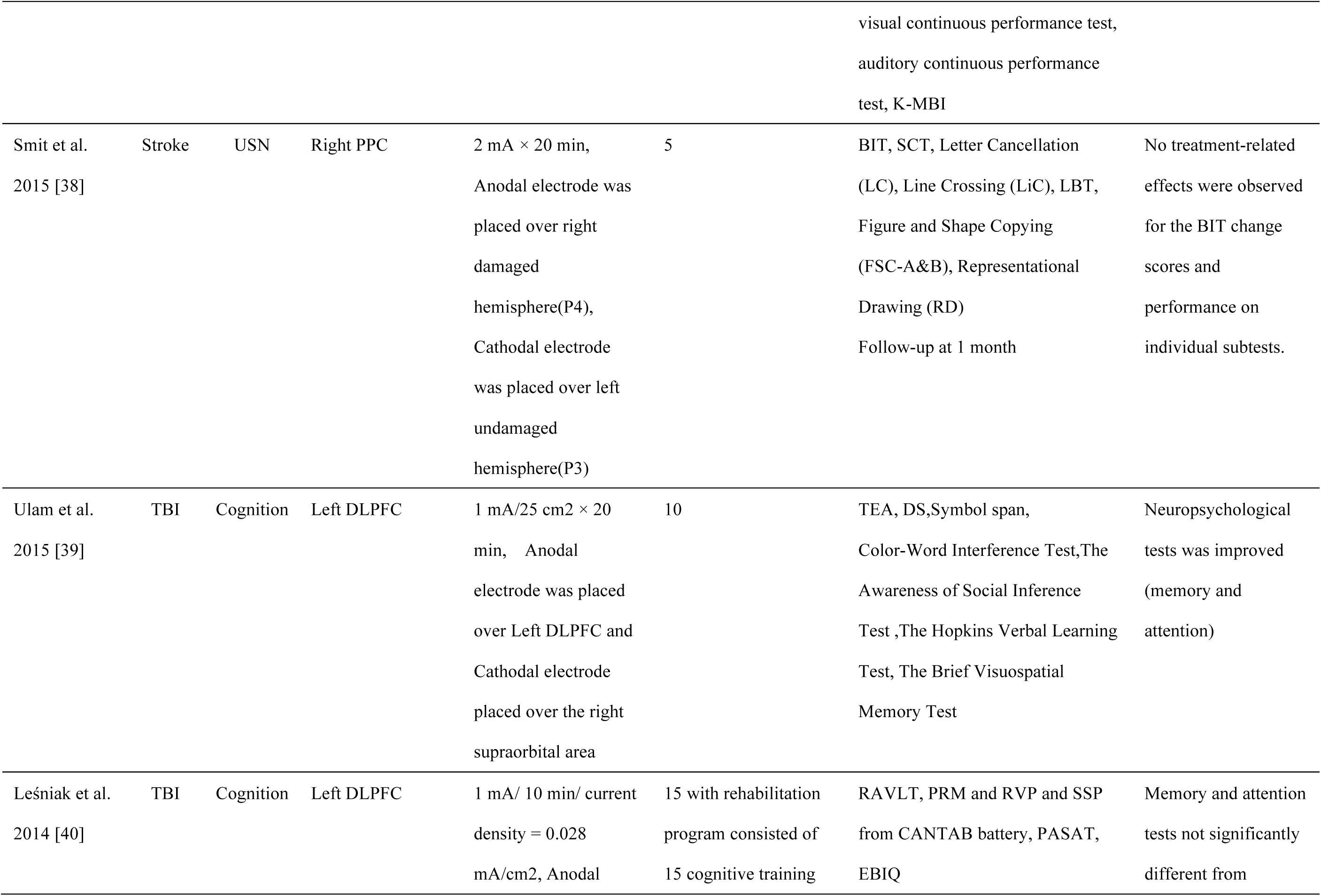
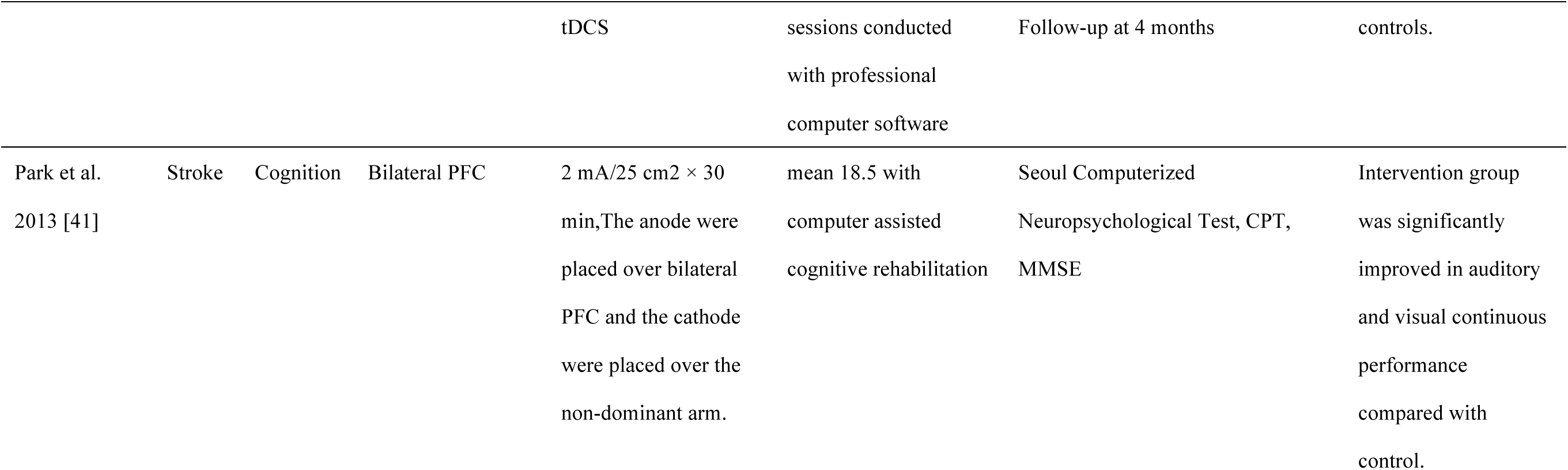
Individual Study Treatment Characteristics, Assessments and Outcomes

| Study | Disease | Targets | Stimulation Site | Parameter | Session | Assessments &<br>Follow-Up | Results |
| --- | --- | --- | --- | --- | --- | --- | --- |
| TMS |  |  |  |  |  |  |  |
| Kim et al.<br>2018 [25] | Stroke | USN | Left PPC | 0.9Hz 95%MT 900<br>pulses/session | 10(rTMS only, Robot<br>only, combined) All<br>three groups received<br>conventional<br>rehabilitation | MVPT-3, LBT, SCT, Albert's test,<br>CBS, MMSE, K-MBI. | There was no<br>significant differences<br>among group. |
| Lee et al.<br>2018 [26] | TBI | Cognition | Right DLPFC | 1Hz 100%MT 2000<br>pulses/session | 10(All patients<br>received<br>neurodevelopmental<br>therapy) | MARDS,TMT, SCWT | Cognition was<br>significantly<br>improved. |
| Cha et al.<br>2016 [27] | Stroke | USN | Left PPC | 1Hz 90%MT 1200<br>pulses/session | 20 with conventional<br>rehabilitation<br>therapy(30 min) | LBT, Albert test, BBT, Grip<br>strength | Intervention group<br>was significantly<br>improved |
| Yang et al.<br>2015 [28] | Stroke | USN | Left PPC | 1Hz 80%MT 656<br>pulses/session, 10Hz<br>80% MT 1000<br>pulses/session,<br>cTBS30Hz 80% MT<br>801 pulses, in bursts of 3 pulses repeated<br>every 100 ms/session | 20 with speech training<br>for 3–4 h per day, 5<br>days a week for a total<br>8 weeks of treatment. | SCT, LBT<br>Follow-up at 1 month | All intervention group<br>was improved<br>compared than Sham<br>group. |
| Lu et al. 2015<br>[29] | Stroke | Memory | Right DLPFC | 1Hz 100%MT 600<br>pulses/session | 20 and received regular<br>computer-assisted<br>cognitive training for<br>30 min every day. | MoCA, LOTCA, RBMT<br>Follow-up at 3 days and 2 months | RMBT was improved.<br>MoCA, LOTCA was<br>not. |
| Kim et al.<br>2010 [30] | Stroke | Cognition | Left DLPFC | 1Hz 900 pulses/ 10Hz<br>450 pulses 80% MT | 10(all participants<br>received conventional<br>cognitive rehabilitation<br>two or three times a<br>week for 2 wks) | A digit span test (forward and<br>backward), visual span test<br>(forward and backward), verbal<br>learning test, visual learning test,<br>visual continuous performance test,<br>auditory continuous performance<br>test, and a word-color test ,Tower<br>of London test, Modified Barthel<br>Index, Beck Depression Inventory | There was no<br>significant<br>improvement about<br>cognition. |
| **TBS** |  |  |  |  |  |  |  |
| Fu et al. 2015<br>[31] | Stroke | USN | Left PPC | 30Hz cTBS repeated<br>every 200 ms 40 s<br>4trails interval 15min | 14(All patients<br>received conventional<br>rehabilitation training,<br>based on visuospatial<br>scanning, of 30 min<br>twice daily for 2 | SCT, LBT<br>Follow-up at 4 weeks | SCT and LBT was<br>improved at 4weeks |

|  |  |  |  |  | weeks. Patients were<br>also treated with<br>programmes for motor<br>rehabilitation when<br>necessary.) |  |  |
| --- | --- | --- | --- | --- | --- | --- | --- |
| tDCS |  |  |  |  |  |  |  |
| Shaker et al.<br>2018 [32] | Stroke | Cognition | Bilateral DLPFC | 2 mA × 30 min, The<br>cathode was placed<br>over the contralateral<br>supraorbital area. | 12 in combination with<br>selected cognitive<br>training program by<br>RehaCom. | Computer-based cognitive therapy<br>tool, FIM | There was a<br>significant<br>improvement in the<br>scores of attention and<br>concentration, figural<br>memory, logical<br>reasoning, reaction<br>behavior. |
| Hosseinzadeh<br>et al. 2018<br>[33] | Stroke | Attention | anoda :left STG,<br>cathoda: Right<br>PPC | 2 mA/35 cm2 × 30<br>min | 12 | NIHSS, TMT, Beck test<br>Follow-up at 1 and 3 months | TMT wasn't<br>significantly changed<br>in all group. NIHSS,<br>Beck test was<br>improved in Anodal. |
| Turgut et al.<br>2018 [34] | Stroke | USN | Left PPC | 1.5-2mA The anode<br>was placed<br>ipsilesional. | 8 during the<br>performance of an<br>optokinetic task and | Apples Cancellation, Spontaneous<br>body orientation, LBT, Cued body<br>orientation, The Clock Drawing | The intervention<br>group improved |
|  |  |  |  |  | received standard<br>therapy for five hours<br>per day. | Test,<br>body orientation, Line Bisection<br>Test, Clock Drawing Test<br>Follow-up at 5-6 days |  |
| Sacco et al.<br>2016 [35] | TBI | Cognition | Bilateral DLPFC | 2 mA/35 cm<sup>2</sup> × 20<br>min, Two anodes, one<br>on the right DLPFC<br>and the other on the<br>left DLPFC, earth on<br>the arm | 10 with<br>computer-assisted<br>training | DA, RBANS, BDI, AES<br>Follow-up at 1 month | The intervention<br>group significantly<br>improved in attention<br>test |
| Yi et al. 2016<br>[36] | Stroke | USN | anodal: right PPC,<br>cathodal: left PPC | 2 mA/25 cm<sup>2</sup> × 30<br>min | 15 with conventional<br>occupational therapy | MVPT,SCT,LBT,CBS,K-MBI,FAC | Improvements in the<br>MVPT, SCT, and<br>LBT were greater in<br>the anodal and<br>cathodal groups than<br>in the sham group. |
| Yun et al.<br>2015 [37] | Stroke | Cognition | fronto-temporal(T3<br>or T4) | 2 mA/25 cm<sup>2</sup> × 30<br>min The anode was<br>placed over T3 or T4. | 15 with cognitive<br>rehabilitation | Korean-MMSE, Computerized<br>neurocognitive function tests,A<br>digit span test (forward and<br>backward), visual span test<br>(forward and backward),verbal<br>learning test, visual learning test, visual continuous performance test,<br>auditory continuous performance<br>test, K-MBI | Left anodal tDCS<br>improved backward<br>digit span task and<br>verbal memory, no<br>improvements in<br>other cognitive tasks. |
| Smit et al.<br>2015 [38] | Stroke | USN | Right PPC | 2 mA × 20 min,<br>Anodal electrode was<br>placed over right<br>damaged<br>hemisphere(P4),<br>Cathodal electrode<br>was placed over left<br>undamaged<br>hemisphere(P3) | 5 | BIT, SCT, Letter Cancellation<br>(LC), Line Crossing (LiC), LBT,<br>Figure and Shape Copying<br>(FSC-A&B), Representational<br>Drawing (RD)<br>Follow-up at 1 month | No treatment-related<br>effects were observed<br>for the BIT change<br>scores and<br>performance on<br>individual subtests. |
| Ulam et al.<br>2015 [39] | TBI | Cognition | Left DLPFC | 1 mA/25 cm<sup>2</sup> × 20<br>min, Anodal<br>electrode was placed<br>over Left DLPFC and<br>Cathodal electrode<br>placed over the right<br>supraorbital area | 10 | TEA, DS,Symbol span,<br>Color-Word Interference Test,The<br>Awareness of Social Inference<br>Test ,The Hopkins Verbal Learning<br>Test, The Brief Visuospatial<br>Memory Test | Neuropsychological<br>tests was improved<br>(memory and<br>attention) |
| Leśniak et al.<br>2014 [40] | TBI | Cognition | Left DLPFC | 1 mA/ 10 min/ current<br>density = 0.028<br>mA/cm<sup>2</sup>, Anodal | 15 with rehabilitation<br>program consisted of<br>15 cognitive training | RAVLT, PRM and RVP and SSP<br>from CANTAB battery, PASAT,<br>EBIQ | Memory and attention<br>tests not significantly<br>different from |

|  |  |  |  | tDCS | sessions conducted<br>with professional<br>computer software | Follow-up at 4 months | controls. |
| --- | --- | --- | --- | --- | --- | --- | --- |
| Park et al.<br>2013 [41] | Stroke | Cognition | Bilateral PFC | 2 mA/25 cm<sup>2</sup> × 30<br>min, The anode were<br>placed over bilateral<br>PFC and the cathode<br>were placed over the<br>non-dominant arm. | mean 18.5 with<br>computer assisted<br>cognitive rehabilitation | Seoul Computerized<br>Neuropsychological Test, CPT,<br>MMSE | Intervention group<br>was significantly<br>improved in auditory<br>and visual continuous<br>performance<br>compared with<br>control. |

### Outcomes

#### Effect of rTMS

Figure 2 shows the relationship between the stimulation site and the neuropsychological test results herein. Two out of seven studies using rTMS reported on global cognition as an outcome, one on memory, and four on USN. In studies reporting on global cognition, Lee et al.^26^ used 1 Hz (inhibitory) stimulation to the right dorsolateral prefrontal cortex (DLPFC). They reported significant improvements in the Trail making test (TMT) and The Stroop Color Word Test (SCWT). On the other hand, Kim et al.^30^ used 1 Hz (inhibitory) or 10 Hz (excitatory) stimulation on the left DLPFC. Neither groups showed any improvement in any neuropsychological tests for memory, attention, and executive function. However, mood state significantly improved with 10 Hz stimulation. Lu et al.^29^ reported that 1 Hz stimulation applied to the right DLPFC improved memory on Rivermead Behavior Memory Test. In the latter study, changes in brain-derived neurotropic factor (BDNF) were examined; BDNF decreased in the intervention group but it increased in the sham group. This change did not correlate with improvements in memory and general cognitive function. In terms of USN, the left posterior parietal cortex (PPC) was selected as the stimulation site in all studies. Inhibitory stimuli were performed in all studies, including one study that selected multiple stimulus patterns. Cha et al.^27^ used 1Hz (inhibitory) stimulation to PPC and Fu et al. ^31^ used 30Hz cTBS stimulation to PPC. Both studies reported that the intervention group showed a significant improvement compared to the control group. Yang et al.^28^ showed significant improvements in all stimulation patterns of 1 Hz, 10 Hz, and cTBS. On the other hand, only the cTBS group showed significant increase in fractional anisotropy (FA) and mean diffusivity (MD) in superior longitudinal fasciculus, superior occipitofrontal fascicle and inferior fronto-occipital fasciculus on the left side (stimulation side) and the capsula external and inferior fronto-occipital fasciculus on the right side (contra-stimulation side). In a study by Kim et al.^25^ using both robotic therapy and rTMS, significant improvements were observed in all groups, but there were no significant differences between the groups.

**Figure 2.**
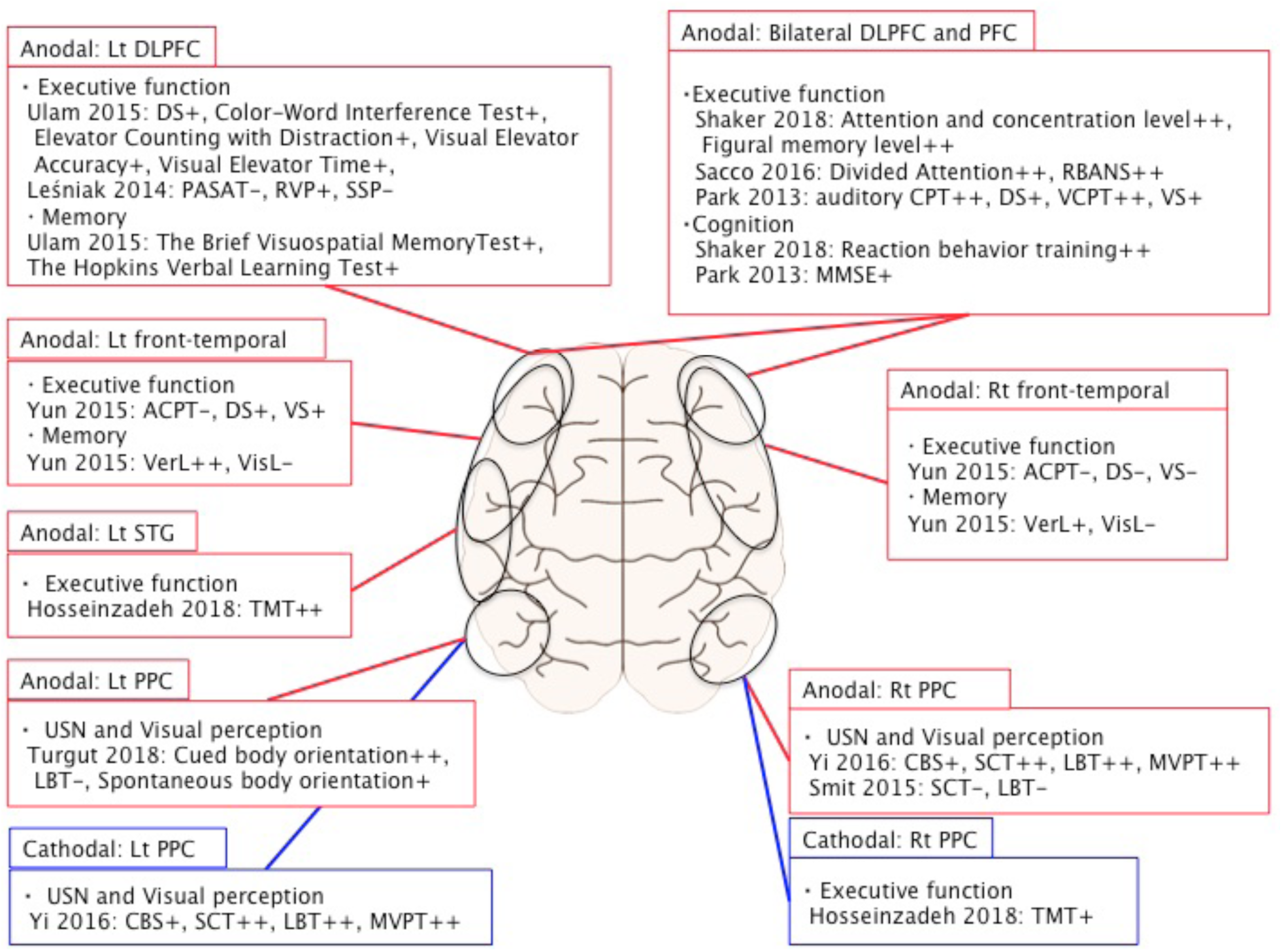
Relationship between stimulation site and neuropsychological test results in tDCS: *Note:* ++ = a significant improvement before and after the intervention and a significant improvement compared to the control group. + = significant improvement before and after the intervention; however, no significant improvement compared to the control group. - = no significant improvement before and after the intervention. *Abbreviations:* DLPFC=Dorsolateral prefrontal cortex, PFC=Prefrontal cortex, PPC=Posterior parietal cortex, STG=Superior temporal gyrus, ACPT=Auditory continuous performance test, CBS=Catherine Bergego Scale, CPT=continuous performance test, DS=Digit Span, LBT= Line bisection test, MMSE=Mini-Mental State Examination MVPT=Motor-Free Visual Perception Test, PASAT=Paced Auditory Serial Addition Test, RBANS=Repeatable Battery for the Assessment of the Neuropsychological Status, RVP=Rapid Visual Processing, SCT=Star cancellation test, SSP=Spatial Span test, TMT=Trail Making Test, VCPT=visual continuous performance test, VerL=verbal learning test, VisL=visual learning test, VS=Visual span

#### Effect of tDCS

Figure 3 shows a summary of the relationship between the stimulation site and the neuropsychological test results described herein. Regarding tDCS, six trials assessed global cognition as the outcome, one assessed attention, and three assessed USN. For global cognition, two trials used bilateral DLPFC as the stimulation site, one trial used the bilateral prefrontal cortex (PFC), two trials used the left DLPFC, and one trial used the fronto-temporal (right or left). Anodal tDCS pattern was used in reports that stimulated the PFC region bilaterally. Shaker et al.^32^ and Sacco et al.^35^ reported significant improvements in executive functions, including attention and memory, and cognitive functions. In addition, Sacco et al.^35^ examined functional Magnetic Resonance Imaging before and after the intervention during divided attention task and showed that brain activity was decreased in the right superior temporal gyrus (BA 42), right and left middle frontal gyrus (BA 6), right postcentral gyrus (BA 3) and left inferior frontal gyrus (BA 9). They indicated that such neural changes were normalization of previously abnormal hyperactivations. Similarly, in a study targeting bilateral PFC, Park et al.^41^ reported that while Digit Span, Visual Span, and MMSE were not significantly improved compared to the control group, the change ratio in auditory and visual continuous performance test that related to sustained and selective attention was significant in the intervention group^41^. In studies that used Anodal tDCS on the left DLPFC, Ulam et al. reported that improvements in attention and working memory tests were observed; however, these were also not significant compared to the control group despite that the improvement correlated with a decrease in delta waves by electroencephalogram measurements^39^. Leśniak et al.^40^ reported improvement for the intervention group in tests for sustained attention and selective attention, although they were not significant compared to the control group. Yun et al.^37^ performed anodal tDCS on the left and right fronto-temporal lobe, compared with sham stimulation, and observed improvement in auditory and visual memory in the left fronto-temporal stimulation; however, only the verbal learning test was significantly improved compared with the control group.

Hosseinzadeh et al.^33^ examined changes in attention function and found that anodal tDCS to the leftSTG had a significant effect compared to the control group. Regarding USN, Smit et al.^38^ reported that there was no significant improvement after performing anodal tDCS on right PPC. On the other hand, Yi et al.^36^ indicated that there was a significant improvement compared to the control group with the same stimulation pattern as described above, and that cathodal tDCS for left PPC also had a significant improvement compared to the control group. Finally, Turgut et al.^34^ found that there was a significant improvement in egocentric neglect using anodal tDCS combined with optokinetic task and rehabilitation for left PPC.

### Safety

Among 17 included studies, 14 reported no obvious side effects. Several studies have reported minor adverse effects. Lu et al.^29^ reported that one patient experienced transient headaches and dizziness in the intervention group. Leśniak et al.^40^ reported that Some patients experienced tingling (n = 6), itching (n = 4), drowsiness (n = 2), headache (n = 1), stinging (n = 1), or dizziness (n = 1); additionally, one patient experienced a panic attack and was consequently excluded from the intervention. Park et al. [41] reported that some patients had pricking sensation (unknown number) at the sites of stimulation after the tDCS.

## DISCUSSION

We performed a systematic review of the effect of NIBS on cognitive impairment for post stroke and TBI. We found that the evidence for positive effects on cognitive function including attention and memory is still limited with significant variability in stimulation target and stimulation parameters. There was significant evidence for a positive effect of NIBS on USN.

There have been some published reviews related to NIBS effect on cognitive impairment; however, overall the evidence for the effect of NIBS for cognitive impairments is still limited. In a systematic review of NIBS for Alzheimer Disease (AD), Hsu et al.^42^ performed a meta-analysis of 11 articles involving 200 patients. The effect of NIBS on cognitive impairment (measured via neuropsychological tests) showed a significant mean effect size of 1.35 (p<0.001 95% CI, 0.86-1.84). Additionally in subgroup analyses, high frequency rTMS stimulation (HFS) showed a significant mean effect size of 1.64 (p<0.001 95% CI, 1.03-2.27) compared to low frequency rTMS stimulation (LFS).^42^ A meta-analysis of seven studies (94 patients) by Liao^43^ examined the effect of rTMS on cognitive impairment in AD. They reported a standard mean difference (SMD) of 1.00 (p<0.001, 95% CI, 0.41-1.58) on rTMS treatment^43^; in subgroup analyses, HFS for right or bilateral DLPFC significantly improved the cognition (SMD=1.06 95%, CI, 0.47-1.66 p<0.05). Lawrence et al. ^44^ report that there is a lack of sufficient evidence to suggest rTMS adequately improves cognition in Parkinson’s Disease (PD). In a review of three articles, the effects of rTMS on executive and attention functions, measured by neuropsychological tests, did not significant improve after rTMS (memory: Hedge’s *g*=0.40 95%, CI, -0.14-0.93 Z=1.46 p>0.05, attention: Hedge’s *g*=0.34 95%, CI, -0.42-1.11 Z=0.88 p>0.05). A review of rTMS on cognition in mild cognitive impairment (MCI) pointed out the possibility that HFS has a better effect than LFS, although the effect size in all seven included studies was small at SMD = 0.48 (p= 0.01 95%CI, 0.12-0.84).^45^ In general, it appears the evidence is limited in several clinical populations.

Many reviews indicate that the region and patterning of excitatory stimulation, particularly centered on the frontal lobe, may be the key observing improvements in cognitive impairment. In our systematic review, we found that anodal tDCS showed improvement for the region centered on the frontal lobe. On the other hand, in rTMS, there was only one report of HFS for the region centering on the frontal lobe. In terms of USN, it was considered effective to stimulate patterns that regulate the imbalance of intercerebral hemisphere inhibition after brain injury.^46^ The frontal lobe and DLPFC are said to be involved in executive function, memory, working memory, and attention.^47-50^ In examining improvement of cognitive function for PD, Dinkelbach suggested that it was effective to select DLPFC as the stimulation site for both rTMS and tDCS, HFS was effective in rTMS, and Anodal tDCS was effective in tDCS.^51^ Therefore, as the report of NIBS for cognitive impairment in AD and PD, these findings suggest that excitatory stimulation pattern, particularly in the frontal lobe, may be effective post-stroke and TBI, but the evidence is still limited. In other diseases, the possibility that HFS for the bilateral frontal region is effective has been reported, and the possibility that the multiple target method is effective has been proposed.^52-54^ Therefore, it is necessary to study these new stimulation parameters and methods for NIBS for cognitive dysfunction after stroke and TBI in future trials.

This review found that few studies reported minor adverse events. The most concerning adverse event was a seizure after rTMS^16^ and seizure and skin burn after tDCS.^55^ No major adverse event was observed in the current review and no studies reported cognitive deterioration after NIBS. To establish routine use of NIBS for cognition post stroke and TBI, it is necessary to establish a method for identifying the lowest-risk stimulation sites and stimulation parameters. Regarding rTMS after TBI, Li et al.^56^ recommend the following from the safety point of view: (a) improved accuracy of the stimulation target using a navigation system and (b) the use of low frequency rTMS. The navigation system can accurately identify the stimulation site and, in addition, can reduce the propagation of the stimulation to the opposing brain function region. In particular, for patients with large brain lesions, precise setting of the stimulation site by the navigation system may be necessary.^57^ LFS can reduce major adverse events such as seizure as compared to HFS. Recently, there have been reports of NIBS using a navigation system after TBI.^21, 58^ Nielson et al^58^ administered rTMS to a patient with depression who had titanium skull plates inserted following surgery for TBI and reported its efficacy and safety.^58^ It is important to note that these case reports utilized LFS, not the previously-described excitatory stimulation pattern in the frontal lobe, which are thought to be effective in improving cognitive function after stroke and TBI. It is indicated that more clinical evidence is needed in the future regarding the relationship between safety and stimulation parameters to improve the effectiveness of treatment. To avoid severe side effects when applying excitatory stimulation, it is necessary to consider not only the navigation system described above, but also the use of medication to reduce stimulation threshold, and monitoring of brain imaging via electroencephalogram.

In this systematic review, cognitive rehabilitation and supplementary cognitive training were conducted in all seven rTMS studies and seven out of ten tDCS studies, of which 78.5% showed improvement in the results of the study. According to previous reports, NIBS in combination with rehabilitation has demonstrated significant improvements in physical functioning and aphasia after stroke and TBI.^46,59^ Restoring impaired neural networks following brain injury is a viable means of promoting functional recovery, and in such a situation, indicates that a strategy to promote network-related reorganization in the brain must be adopted.^60^ NIBS may be a promising complementary treatment when used in conjunction with conventional therapies to enhance rehabilitation in patients with brain injury.^20^ From the concept of rehabilitation aimed at improving neuroplasticity, NIBS combined with rehabilitation suggests the possibility of inducing a positive synergistic effect. In addition, this is thought to lead to, not only modulation of neural connections, but also functional relearning.

Based on this systematic review and our previous studies, to build evidence of NIBS for cognitive dysfunction after stroke and TBI, it is important not only to evaluate neuropsychological tests, but also to establish evidence for the effects of NIBS itself on neural networks. Regarding the effects of NIBS on neural networks, the mechanism of action differs between rTMS and tDCS, and the mechanism of action of NIBS itself still remains an important debate.^17,18^ However, consistent is the potential for NIBS to have a positive impact on pathological rhythms in the network after injury or caused by disease. Previous neuroimaging studies have reported that NIBS affects the cerebral cortex directly under the stimulation site or its functional-related brain regions based on neural networks.^46,61^

In this systematic review, changes in brain activation were evaluated using neuroimaging, and neuropsychological tests.^28,35,39^ The common point in all is that a change in activity in the brain region was associated with the stimulation site. These results are consistent with previous NIBS studies for upper extremity and aphasia after stroke and TBI.^46,63,64^ To make the clinical application of NIBS for cognitive impairment more robust, it is necessary to consider that the site of brain injury varies from patient to patient. The results obtained from NIBS may vary and would be reflected in changes in brain activity using neuroimaging along with neuropsychological tests. As indicated previously, NIBS affects not only the cerebral cortex under the stimulation site but also functional-related brain regions based on neural networks. For example, in a recent study of NIBS for aphasia, it was suggested that the stimulation site and parameter is selected depending on how the damaged language region and homologous regions related to language act on the recovery of language function, based on the duration of onset and the results of changes in brain activity by language task.^65^ In terms of the relationship between NIBS and the effect of neural networks, Padmanbhan et al.^65^ reported the relationship between brain function connectivity and depression in post-lesion depression. Lesion locations associated with depression were highly heterogeneous and there was no consistent brain region related to depression. Lesion locations were mapped to a connected brain circuit centered on the left DLPFC; the size of the damaged area alone could predict depression.^65^ This same observation may be applied to the relationship between brain lesion and symptoms in cognitive impairment in post-stroke and TBI. Kreuzer et al.^66^ also described the relationship and neural connectivity between DLPFC and anterior cingulate cortex in a review of NIBS for DLPFC. They suggested that rTMS for DLPFC has the effect on anterior cingulate cortex which is functionally related to DLPFC. Therefore, they argued that preclinical parameter studies combining TMS with neuroimaging are necessary.^67^ Cognitive evaluation along with neuroimaging evaluation will lead to enhanced evidence of the effectiveness and accuracy of NIBS treatment, as well as the exploration of new insights and methods post stroke and TBI.

## CONCLUSIONS

We performed a systematic review of the efficacy of NIBS on cognitive impairment post-stroke and TBI and found limited evidence for improvements in attention and memory. A significant improvement was observed for USN after NIBS. However, the most optimal stimulation sites and stimulation parameters remain unknown. Based on the stimulation sites and parameters from previous published studies, we conclude that excitability stimulation for the region centered on the frontal lobe is worth studying in the future. In addition, it is suggested that the neural plasticity change induced by NIBS may contribute to greater improvements when combined with rehabilitation. Finally, evaluation of brain activity at the stimulation site and related areas using neuroimaging on how NIBS acts on the neural network will contribute to the establishment of evidence.

## Data Availability

all data are included in the manuscript

## Acknowledgements

**None**

